# Quantifying and adjusting for confounding from health-seeking behaviour and healthcare access in observational research

**DOI:** 10.1101/2024.02.27.24303434

**Authors:** S. Graham, J.L. Walker, N. Andrews, W.J. Hulme, D. Nitsch, E.P.K. Parker, H.I. McDonald

**Affiliations:** Faculty of Epidemiology and Population Health, London School of Hygiene and Tropical Medicine, London, United Kingdom; National Institute for Health and Care Research Health Protection Research Unit in Vaccines and Immunisation, London United Kingdom; UK Health Security Agency, London, United Kingdom; The Bennett Institute for Applied Data Science, Nuffield Department of Primary Care Health Sciences, University of Oxford, Oxford, United Kingdom; UK Renal Registry, Bristol, United Kingdom; Renal Unit, Royal Free London NHS Foundation Trust, Hertfordshire, United Kingdom; Faculty of Science, University of Bath, Bath, United Kingdom

## Abstract

**Objective:** To assess the feasibility and effect of using proxy markers of health-seeking behaviour and healthcare access to quantify and adjust for confounding in observational studies of influenza and COVID-19 vaccine effectiveness (VE).

**Design:** Cohort study for influenza VE in the 2019/2020 influenza season and for early COVID-19 VE (December 2020 – March 2021).

**Setting:** Primary care data pre-linked to secondary care and death data in England.

**Participants:** Individuals aged ≥66 years on 1 September 2019.

**Interventions:** Vaccination with any influenza vaccination in the 2019/2020 season or with either a BNT162b2 or ChAdOx1-S vaccination from 08/12/2020 to 31/03/2021.

**Main outcome measures:** Influenza or COVID-19 specific infections, hospitalisation and death. VE was estimated with sequential adjustment for demographics, underlying health conditions, and 14 markers reflecting uptake of public health interventions (screenings, vaccinations and NHS health checks), active healthcare access/use (prostate antigen testing, bone density scans, GP practice visits, low value procedures and blood pressure measurements) and lack of access/underuse (hospital visits for ambulatory care sensitive conditions and did not attend primary care visits). Influenza vaccination in the 2019/2020 season was also considered as a negative exposure intervention against the first wave of COVID-19.

**Results:** We included 1,991,284, 1,796,667, and 1,946,943 individuals in the influenza, COVID-19 and negative exposure VE populations, respectively. Vaccinated individuals were more likely to display active health-seeking behaviour, including participation in UK national screening programmes, compared with unvaccinated individuals. In the 2019/2020 influenza season, adjusting for health-seeking markers increased VE against infection from −1.5% (95%CI: −3.2,0.1) to 7.1% (95%CI: 5.4,8.7), but this trend was less apparent for more severe outcomes. For COVID-19 during early vaccine roll out, adjusting for health-seeking markers in addition to demographics and underlying health conditions did not change VE estimates against infection or severe disease (e.g., two doses of BNT162b2 against infection: from 82.8% [95%CI: 78.4,86.3] to 83.1% [95%CI: 78.7,86.5]). Adjusting for health-seeking markers removed bias in the negative exposure analysis of influenza vaccination against SARS-CoV-2 infection (−7.5% [95%CI: −10.6,-4.5] vs −2.1% [95%CI: −6.0,1.7] before vs after adjusting for health-seeking markers).

**Conclusions:** Markers of health-seeking behaviour and healthcare access can be identified in electronic health records, are associated with vaccination uptake, and can be used to quantify and account for confounding in observational studies.

**What is already known on this topic:** Health-seeking behaviour and healthcare access are recognised confounders in observational studies, but are not directly measurable in electronic health records (EHRs). Previously we systematically identified 14 markers in UK EHRs that reflect different aspects of health-seeking behaviour and healthcare access. We do not know if these markers can be utilised to quantify and account for this type of confounding in observational research using influenza and COVID-19 vaccine effectiveness as examples.

**What this study adds:** This study demonstrated using the proxy markers that confounding from health-seeking behaviour and healthcare access underestimates influenza VE estimates, but has negligible impact on COVID-19 VE estimates during early vaccine roll out. We also demonstrated via a negative exposure analysis that residual confounding can be removed by adjusting for these proxy markers.

## Background

Health-seeking behaviour and healthcare access may be important confounders in observational research. Health-seeking behaviour is defined as seeking care for disease prevention, when asymptomatic or during early symptomatic stages,(1) whereas healthcare access can be defined as the ability to access healthcare services for these purposes.(2) Individuals with active health-seeking behaviour and the ability to easily access healthcare services are generally expected to have favourable clinical outcomes. Previously confounding from health-seeking behaviour and healthcare access has led to overestimates of effectiveness of some preventative therapies in observational research. For example, observational studies of statin use and hip fracture have consistently shown, even in meta-analysed results, to reduce risk by 23%, even though this has not been reflected in clinical trials(3). In cohort studies of influenza vaccine effectiveness (VE), authors have reported reductions in all-cause mortality by 40-50%(4, 5). Findings from these studies have since been disputed as influenza accounts for a maximum of 10% of deaths per year, so even if the vaccination was 100% effective it would only be able to prevent 10% of deaths per year(6) However, authors do not usually systematically attempt to account for this type of confounding, as health-seeking behaviour and healthcare access is typically not directly measurable in routine healthcare data.

Proxy markers identified in electronic health records (EHRs) have been used previously to account for confounding from health-seeking behaviour and healthcare access(7–11). Markers included in these studies vary considerably and therefore the best approach to account for this type of confounding is unclear. We recently identified a systematic set of fourteen markers of health-seeking behaviour and healthcare access in UK EHRs(12) that accounted for a range of determinants from a behavioural model known as the Theory of Planned Behaviour.(13) These markers represented interactions with the healthcare system that are only partially driven by an individual’s underlying health need. In the current study, we aimed to assess whether these markers or proxies of health-seeking behaviour and healthcare access in UK EHRs data can be used to quantify and adjust for confounding in observational studies, using seasonal influenza and COVID-19 VE as examples.

## Methods

### Data sources

We conducted a cohort study using Clinical Practice Research Datalink (CPRD) Aurum(14) data pre-linked to Hospital Episode Statistics (HES) Admitted Patient Care (APC)(15) and Office for National Statistics (ONS) death and socioeconomic status data.(16) CPRD Aurum includes diagnoses, prescriptions, referral and testing information of patients registered to consenting GP practices in the UK that use EMIS® software systems. Medical diagnoses are recorded using SNOMED, Read Coded Clinical Terms version 3 (CTV3), or local EMIS® codes that are each mapped to a unique medcode. Information on prescription medicines is coded using the Dictionary of Medicines and Devices (dm+d) codes that are each mapped to a unique prodcode. CPRD Aurum data was pre-linked to HES APC, which includes all admissions to NHS hospitals in England(15). It includes inpatient hospital admission and discharge dates, diagnoses recorded using International Classification of Diseases 10th (ICD-10) Revision codes(17) and procedures recorded using Operating Procedure Codes Supplement (OPCS) codes(18). The data was also pre-linked to ONS, which includes date and underlying cause of death, recorded using ICD-10 codes, and socioeconomic data based on multiple deprivation index(19) which is based on small area geographical location. Linkage from CPRD Aurum to HES APC and ONS datasets occurs at the patient level and is consented at practice level. At the time of data extraction (May 2022 release), the CPRD Aurum data included 1,345 general practices for 13,300,067 currently contributing patients (19.8% of the UK population)(20).

### Study design and population selection

We created separate cohorts to estimate influenza and COVID-19 VE. In addition, to assess for potential residual confounding, we created a third “negative exposure” cohort. Negative exposures assume there to be no causal mechanism between the negative exposure and outcome, but for the confounding structures to reflect those of the primary exposure(21). In the current study we used seasonal influenza vaccinations in 2019/20 as a negative exposure against COVID-19 infections before COVID-19 vaccinations were available in the UK.

An overview of the study design for each of these three populations can be found in Figure 1 and further details in Supplementary Table 1. We included all individuals aged ≥66 years on 1 September 2019. This age group is prioritised for vaccination (including for seasonal influenza vaccine and primary COVID-19 vaccination), and is also likely to show distinct patterns of health-seeking behaviour and healthcare access varies (e.g. due to eligibility of various age-specific screening programmes)(22). We required all individuals to have at least one year of registration prior to their index date (Figure 1 and described further below), have a record of ‘acceptable’ quality by CPRD, and be eligible for linkage to HES APC and ONS. We excluded individuals with a death or registration end date before index date. We also excluded those with indeterminate sex due to low numbers. Individuals in the COVID-19 cohort were additionally excluded if they received their first vaccination prior to 8 December 2020 as these were likely to reflect coding errors or trial participants with a different risk profile.

**Figure 1.**
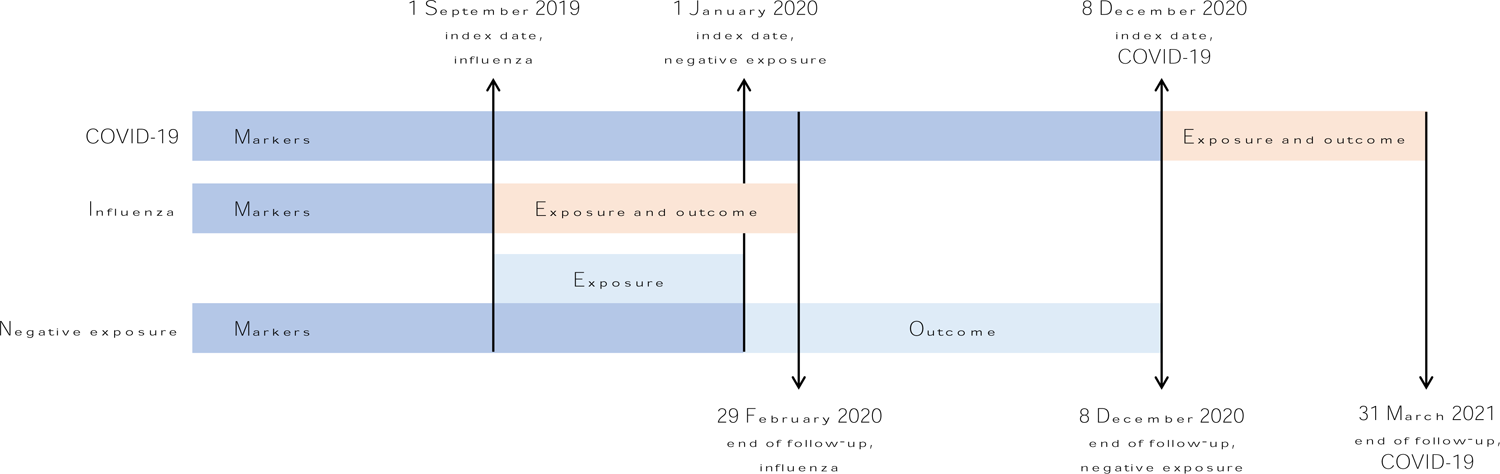
Study design. We followed individuals until the earliest of death, transfer out of GP practice, end of data availability (COVID-19), end of influenza season (influenza), or start of COVID-19 vaccination roll out (negative exposure). For the COVID-19 cohort, we censored individuals at first COVID-19 vaccination that was neither BNT162b2 or ChAdOx1, or on receipt of a second heterologous vaccination.

### Outcomes, exposures, and follow-up

For all analyses we considered three nested outcomes of increasing severity: infections (based on GP data in CPRD Aurum, hospitalisations in HES APC or deaths in ONS); hospitalisation/deaths; and deaths. All COVID-19 outcomes were required to have a COVID-19 diagnosis code. For influenza we required a diagnosis of acute respiratory infection or influenza like illness (ARI/ILI) using code lists from Davidson et al, 2021(23). For all hospital and death outcomes the diagnosis code was required to be in the primary position. Further details on the study outcome and exposure definitions can be found in Supplementary Table 1.

For influenza a first dose of any seasonal influenza vaccination that was recorded during the 2019/2020 season was included. We identified doses of the COVID-19 vaccines BNT162b2 and ChAdOx1 separately. We also identified second doses for COVID-19, requiring a minimum interval of at least 18 days from first to second dose to reflect Joint Committee on Vaccination and Immunisation dosing schedules.(24) Only prodcodes were used to identify COVID-19 vaccinations, as during the COVID-19 pandemic, COVID-19 vaccinations were automatically recorded in patient GP records. For influenza, we identified vaccinations using both medcodes and prodcodes. Since individuals can have more than one medcode recorded on the same day that can represent conflicting information (e.g., given and not given), an algorithm was used to classify influenza vaccination status (see Supplementary Table 1).

For influenza, we used an index date of 1 September 2019. Individuals were followed up from this date until the earliest of death, transfer out of the practice or start of the COVID-19 pandemic (29 February 2020). For COVID-19, we used an index date of 8 December 2020, when COVID-19 vaccinations were first introduced in the UK. Individuals were followed up from this date until the earliest of death, transfer out of the practice, end of data availability (31 March 2021), first vaccination that was neither BNT162b2 or ChAdOx1, or second heterologous vaccination. For the negative exposure analysis, we used an index date of 1 January 2020 as first SARS-CoV-2 infections were identified in the UK after this date. We only included influenza vaccinations before 31 December 2019 by which time the majority of vaccinations in the UK have been delivered(25). This was to reflect positive health-seeking behaviour/access, and prevent overlap with the outcome period, when vaccination uptake might have been influenced in part by awareness and attitudes towards the emerging COVID-19 pandemic. Individuals were followed-up from this date until the earliest of death, transfer out of the practice, or the day before introduction of COVID-19 vaccinations in the UK (7 December 2020).

### Sociodemographic variables at index

At index for each cohort we described the following variables: age (day and month imputed as 01/07 for all individuals as only year of birth is recorded in CPRD), sex, recent infection, local area index of multiple deprivation (IMD), ethnicity, and influenza ‘at-risk’ conditions(26). Recent infection for SARS-CoV-2 was defined as an infection in the three months pre-index, whereas for influenza this was defined as an influenza infection in the previous influenza season. IMD was identified from the ONS at the patient level, or if missing by the primary care practice. Ethnicity was identified from primary care records as described by Mathur et al.(27). Briefly, the algorithm uses a modal approach with ties resolved by recency. If ethnicity could not be identified in primary care, then ethnicity from HES APC was used.

Influenza ‘at-risk’ groups(26) were identified using primary care records of diagnoses or medications as described previously(23), and grouped into immunosuppressed status or other conditions. Individuals with missing age and sex are not provided by CPRD. We assessed missingness of ethnicity, IMD and region. For all other variables, if codes did not appear in an individual’s health record, this was regarded as evidence of absence. Further details on how variables were defined can be found in Supplementary Table 1.

### Markers of health-seeking behaviour and healthcare access at index

We used 14 markers of health-seeking behaviour and healthcare access that we identified through a framework based on the updated Theory of Planned Behaviour(13), as described previously.(12) These included markers representing uptake of public health interventions (AAA, breast cancer, cervical cancer and bowel cancer screening; influenza and pneumococcal vaccination and NHS health checks), active healthcare access/use (prostate antigen testing, bone density scans, GP practice visits, low value procedures and blood pressure measurements) and lack of access/underuse (hospital visits for ambulatory care sensitive conditions and did not attend primary care visits). The markers were identified in CPRD Aurum using medcodes or in HES APC using OPCS or ICD-10 codes as described previously.(12) The lookback periods reflect use of these markers in UK clinical practice and are detailed in Supplementary Table 1.

### Statistical analyses

We described sociodemographic variables, clinical variables, and markers of health-seeking behaviour at index stratified by final vaccination status. To assess timeliness of vaccination, we calculated median days from index to first vaccination, stratified by marker status and age categories (since for COVID-19 vaccinations the UK phased approach was based on age) amongst vaccinated individuals. We grouped the older age categories (over 85 years) due to low patient numbers. Outcome rates were represented by vaccination status as number of events divided by total person years. Cox regression models were used to estimate the risk of infection in vaccinated versus unvaccinated individuals. A complete case analysis was conducted (excluding individuals with a missing value for age, sex, ethnicity or IMD). In the influenza and COVID-19 analyses, vaccination status was time-updated, such that all individuals started follow-up unvaccinated at index date and their vaccination status was updated 14 days after a vaccine was administered (to provide time for immune response induction). For COVID-19, we assessed VE (defined as [1 – hazard ratio] x 100) for BNT162b2 and ChAdOx1 by fitting separate models each restricted to unvaccinated individuals and those who received the vaccine of interest. In the negative exposure analysis, since the vaccination and outcome periods did not overlap, a history of influenza vaccination in the previous season was defined as a binary exposure at index date. We adapted a previous modelling strategy(28) to understand the hierarchical relationships between determinants of vaccine uptake in four iterative steps. First, we fitted baseline models (Model 1) that adjusted for age (as a quadratic polynomial), sex, region and recent infection. In demography-adjusted models (Model 2), we further adjusted for ethnicity and IMD. In comorbidity-adjusted models (Model 3), we further adjusted for immunosuppressive status and other comorbidities. Finally, we fitted fully adjusted models (Model 4) that further adjusted for markers of health-seeking behaviour and healthcare access. For sex-specific markers (cervical cancer screening, breast cancer screening, AAA screening and PSA test), we included an interaction term with sex in the fully adjusted models.

### Sensitivity analysis

We conducted a sensitivity analysis to re-run the fully adjusted models fitting age interactions with AAA screening, bowel cancer screening, NHS health checks and ACS conditions. These were selected as we previously found their prevalence to vary markedly by age.(12)

### Patient and public involvement

There was no patient or public involvement in the study design or interpretation of results.

### Ethics and CPRD requirements

Our study was approved by CPRD’s (#21_000737) and LSHTM’s (#28169) independent ethics review committees. To meet CPRD patient confidentiality requirements we redacted counts relating to less than 5 individuals and conducted secondary suppression where necessary.

### Software, code and reproducibility

All the analyses were conducted using R versions 4.2.2 to 4.2.4. All programming code from this project can be found in the Github repository: https://github.com/grahams99/Health-seeking-behaviour. The code lists and related search terms can be found on LSHTM Data Compass (https://doi.org/10.17037/DATA.00003684).

## Results

### Study population

We included 1,946,943, 1,796,667 and 1,991,284 individuals in the influenza, COVID-19 and negative exposure cohorts, respectively (Figure 2). Compared with individuals who remained unvaccinated throughout enrolment, vaccinated individuals were more likely to be older, of White ethnicity, and live in less deprived areas. Baseline demographic, underlying health conditions, and health-seeking characteristics are summarised for each cohort in Supplementary Table 3, and stratified by subsequent vaccination status in Table 2.

**Figure 2.**
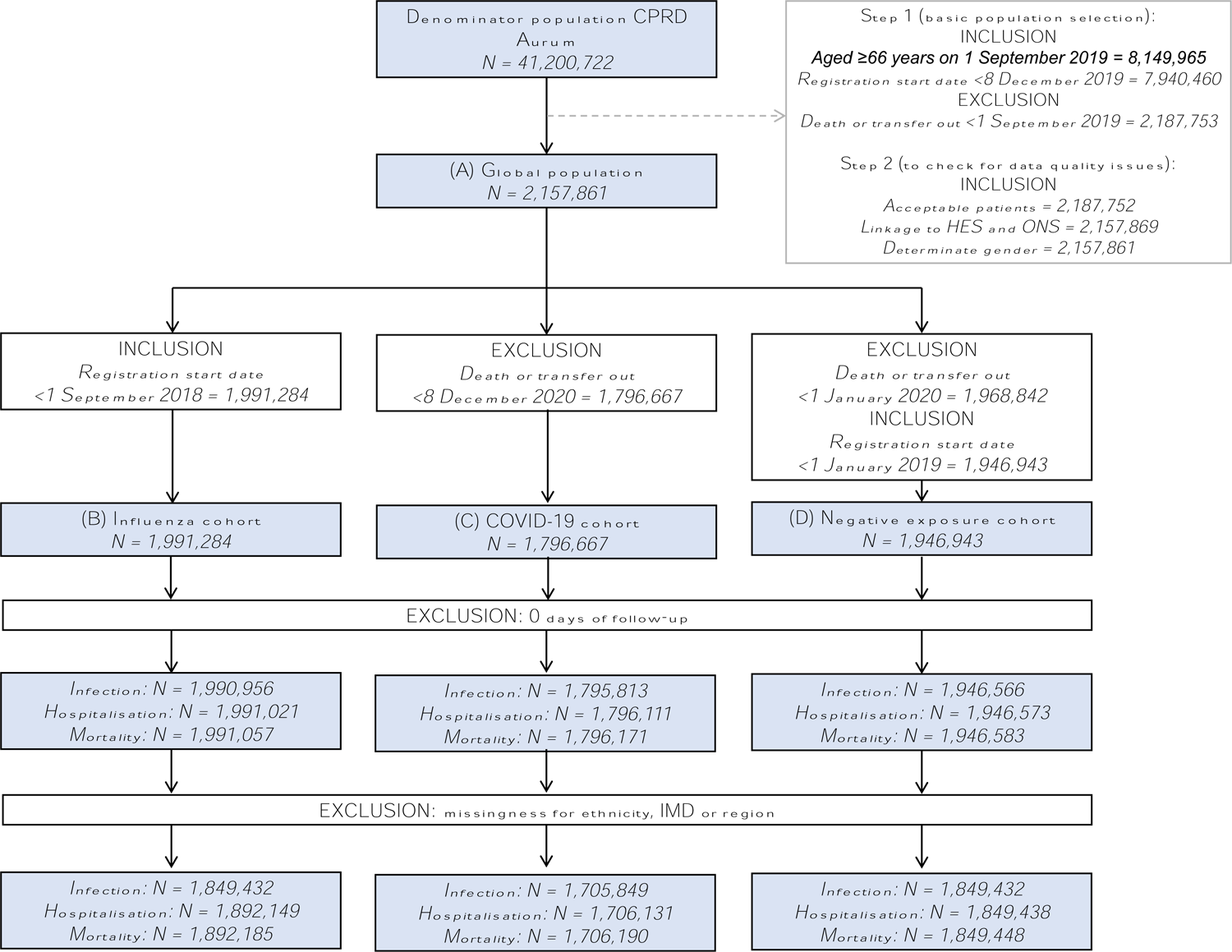
Study selection criteria. Additional details on population selection can be found in Supplementary Table 1. Abbreviations: CPRD: clinical practice research datalink; HES: hospital episode statistics; IMD: index of multiple deprivation; ONS: office for national statistics.

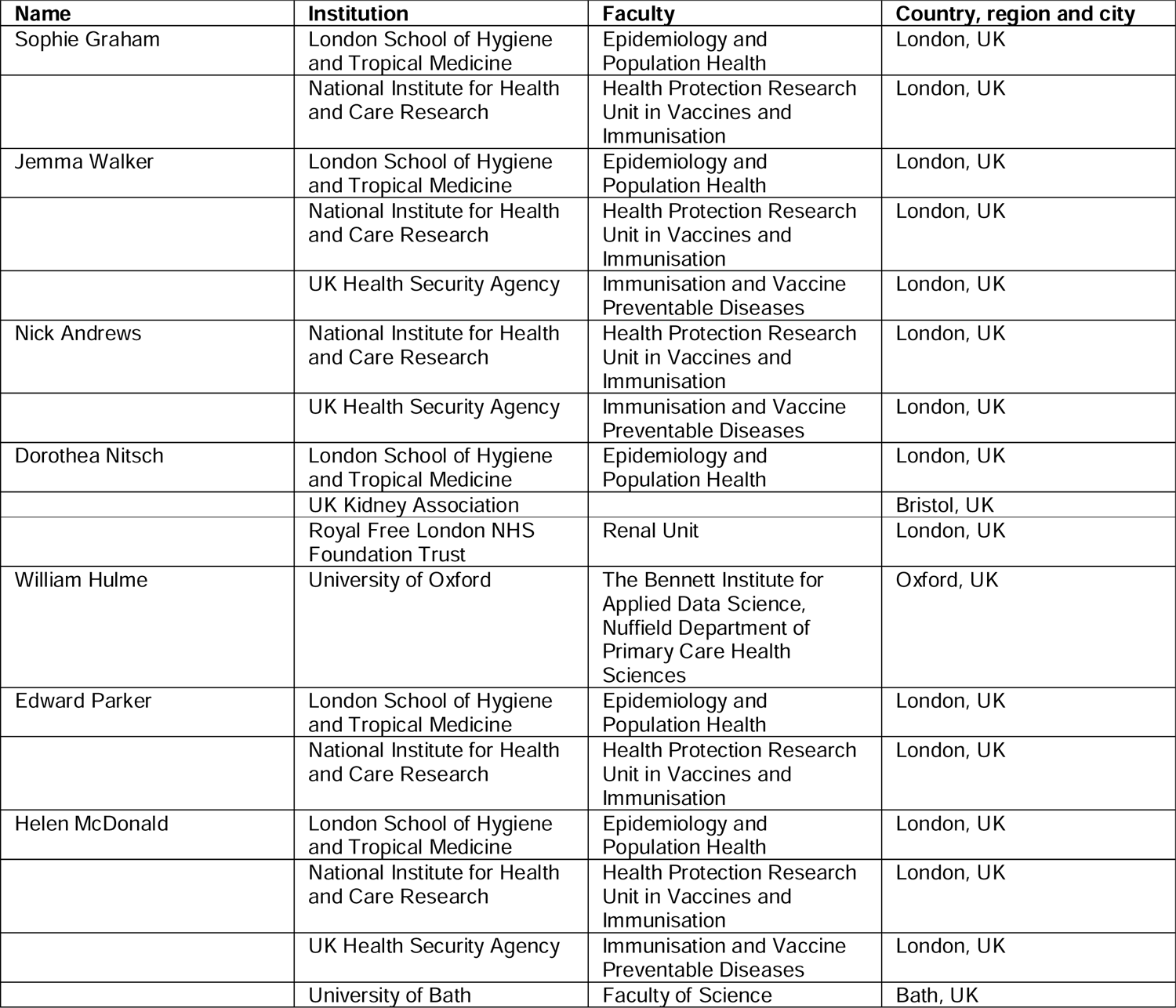

| Name | Institution | Faculty | Country, region and city |
| --- | --- | --- | --- |
| Sophie Graham | London School of Hygiene and Tropical Medicine | Epidemiology and Population Health | London, UK |
|  | National Institute for Health and Care Research | Health Protection Research Unit in Vaccines and Immunisation | London, UK |
| Jemma Walker | London School of Hygiene and Tropical Medicine | Epidemiology and Population Health | London, UK |
|  | National Institute for Health and Care Research | Health Protection Research Unit in Vaccines and Immunisation | London, UK |
|  | UK Health Security Agency | Immunisation and Vaccine Preventable Diseases | London, UK |
| Nick Andrews | National Institute for Health and Care Research | Health Protection Research Unit in Vaccines and Immunisation | London, UK |
|  | UK Health Security Agency | Immunisation and Vaccine Preventable Diseases | London, UK |
| Dorothea Nitsch | London School of Hygiene and Tropical Medicine | Epidemiology and Population Health | London, UK |
|  | UK Kidney Association |  | Bristol, UK |
|  | Royal Free London NHS Foundation Trust | Renal Unit | London, UK |
| William Hulme | University of Oxford | The Bennett Institute for Applied Data Science, Nuffield Department of Primary Care Health Sciences | Oxford, UK |
| Edward Parker | London School of Hygiene and Tropical Medicine | Epidemiology and Population Health | London, UK |
|  | National Institute for Health and Care Research | Health Protection Research Unit in Vaccines and Immunisation | London, UK |
| Helen McDonald | London School of Hygiene and Tropical Medicine | Epidemiology and Population Health | London, UK |
|  | National Institute for Health and Care Research | Health Protection Research Unit in Vaccines and Immunisation | London, UK |
|  | UK Health Security Agency | Immunisation and Vaccine Preventable Diseases | London, UK |
|  | University of Bath | Faculty of Science | Bath, UK |

**Table 2.**
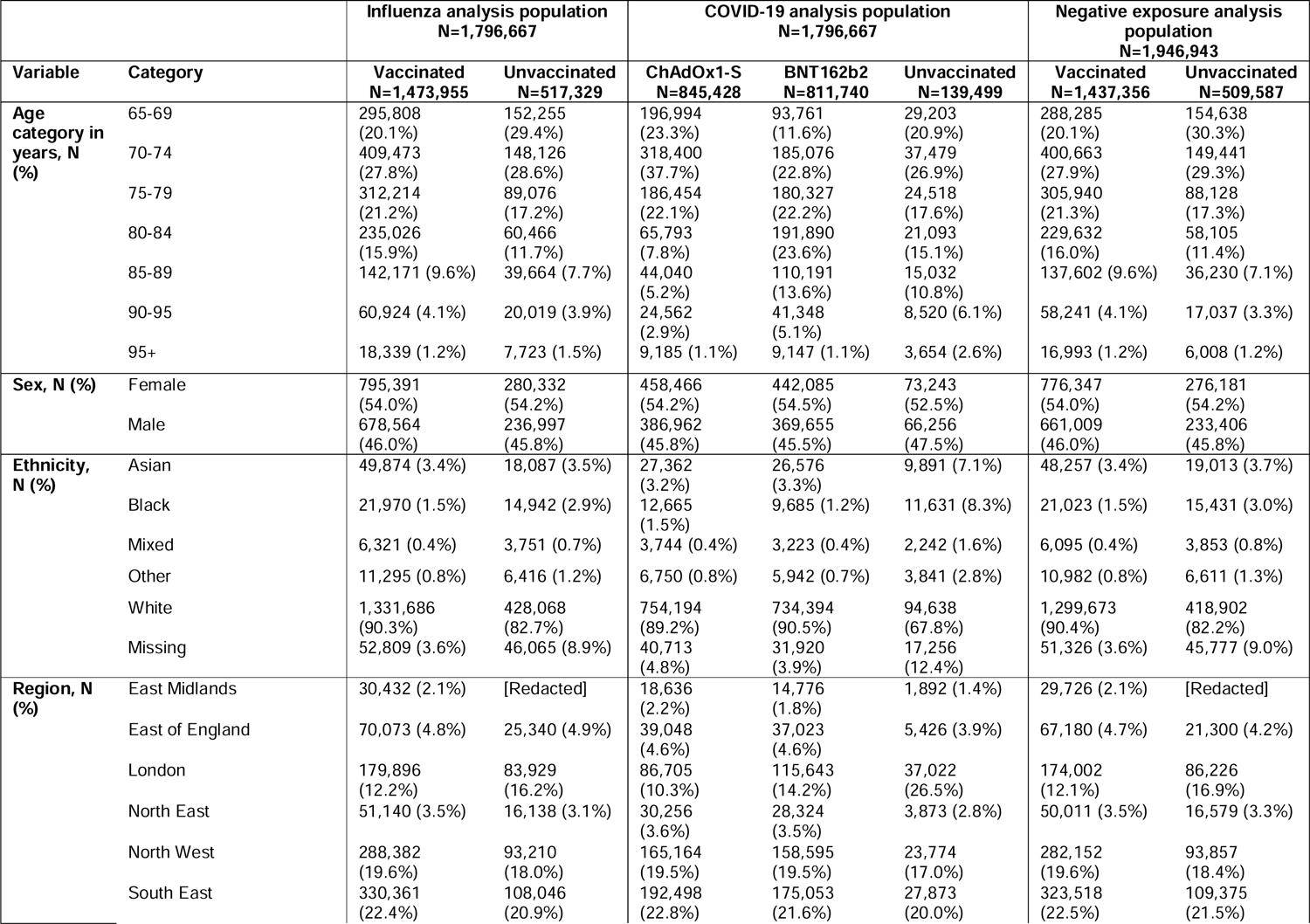
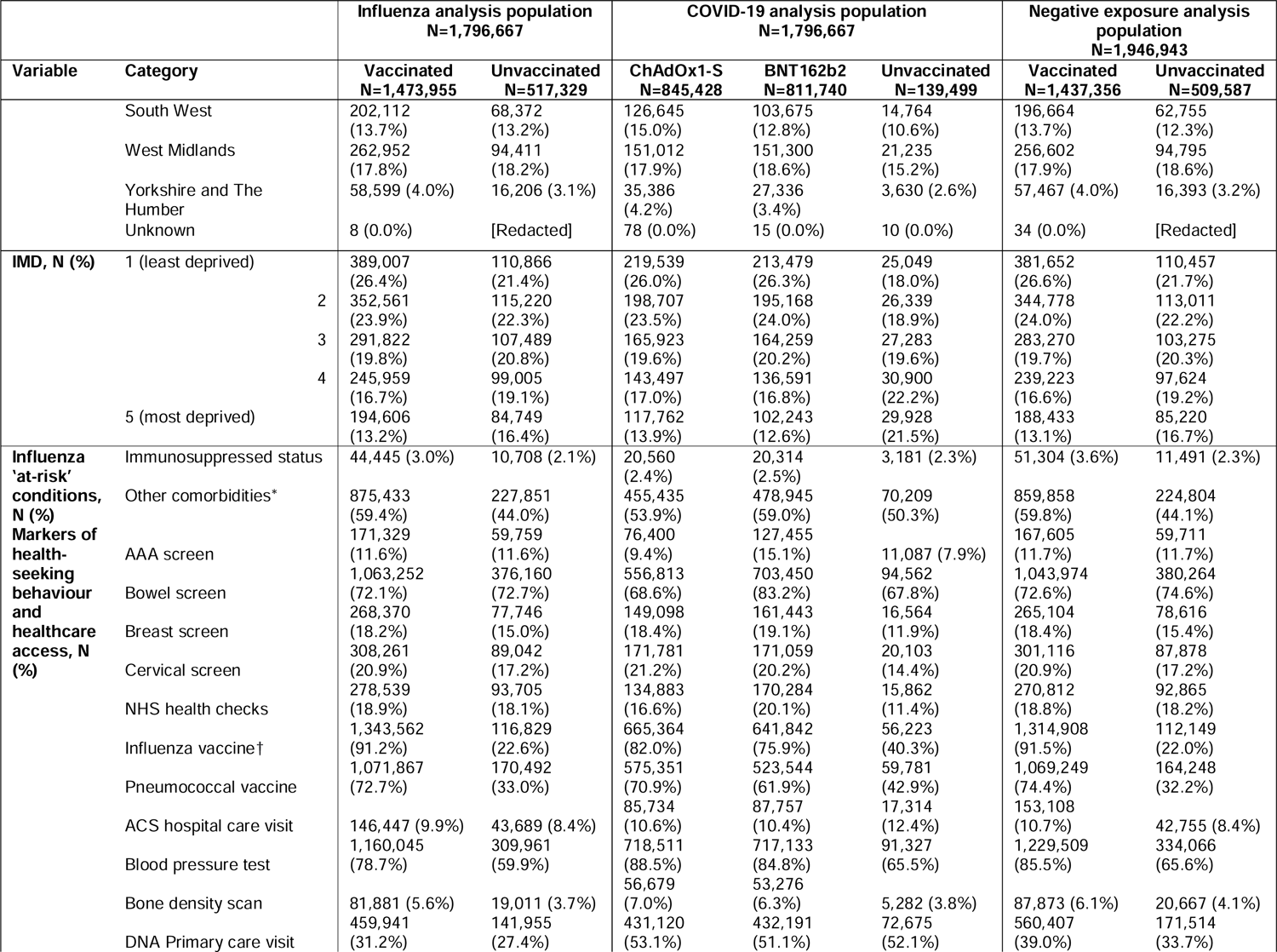
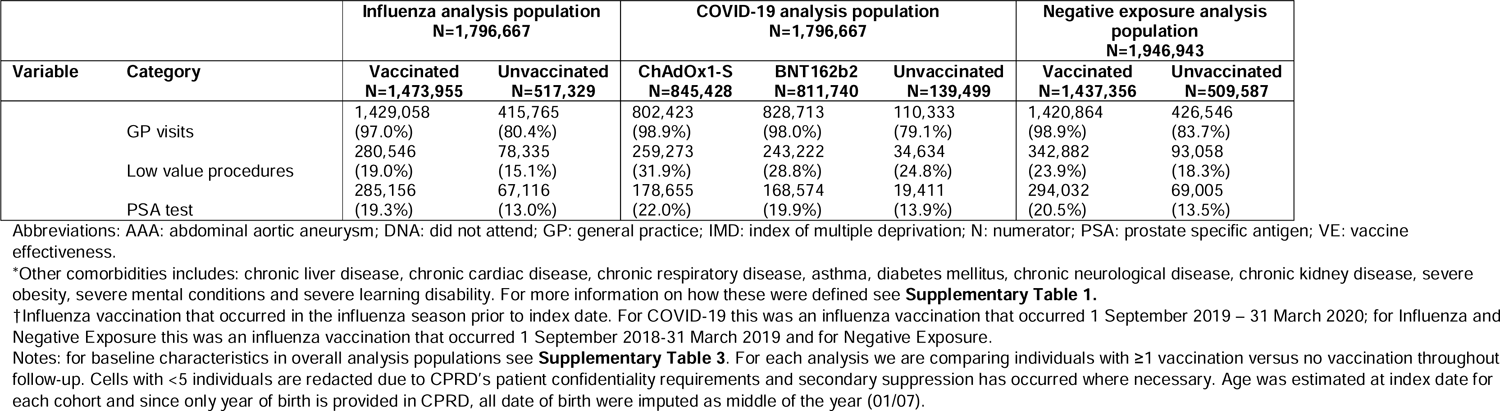
Baseline characteristics stratified by vaccination status at the end of follow-up.

### Markers of health-seeking behaviour

Compared with individuals that remained unvaccinated throughout enrolment, vaccinated individuals had a higher prevalence of all markers of health-seeking behaviour and healthcare access (with the exception of hospital visits for ambulatory care sensitive conditions; Table 2). The differences were greatest for previous vaccinations. In the influenza analysis, 91.2% of vaccinated had an influenza vaccination in the previous season, versus 22.6% in those unvaccinated for influenza vaccination. 72.7% of influenza vaccinated individuals had a pneumococcal vaccination prior to index, compared with only 33.0% in those unvaccinated for influenza. The same was true for the COVID-19 analysis, with 82.0% of ChAdOx1-S vaccinated and 75.9% of BNT162b2 vaccinated individuals having an influenza vaccination in the previous season versus 40.3% in those unvaccinated for any COVID-19 vaccination. 70.9% of individuals vaccinated for ChAdOx1-S and 61.9% of individuals vaccinated for BNT162b2 had a pneumococcal vaccination prior to index, compared with 42.9% in individuals unvaccinated for COVID-19. Among vaccinated individuals, time to vaccination was not strongly associated with health-seeking marker status, with the exception of influenza vaccination in the previous season, which was associated with faster uptake of both COVID-19 and influenza vaccines (Supplementary Figure 1 and 2 with complementary raw data in Supplementary Table 6 and 7).

### Vaccine effectiveness estimates

For the influenza analysis population, median (IQR) follow up time was 181 (0) days. Follow-up time after first influenza vaccination was median of 50 (28) days. Unadjusted event rates ranged from 0.84 per 1,000 person-years during unvaccinated time against ARI/ILI-related death to 117.15 per 1,000 person-years after vaccination against influenza infections (Supplementary Table 4). Incremental adjustments across the models led to increased VE estimates. For influenza infections, we observed a negative VE in the minimally adjusted baseline models of −5.5% (95%CI: −7.2,-3.9). Estimated VE increased to −1.5% (95%CI: −3.2,0.1) after adjusting for comorbidities, and to 7.1% (95%CI: 5.4,8.7) after adjusting for all 14 markers of health-seeking behaviour and healthcare access. For severe outcomes, VE estimates were more consistent across models, ranging from 42.5% (95%CI: 32.8 - 50.8) in the baseline model to 47.5% (95%CI: 37.3 - 56.1) for ARI/ILI-related death in the fully adjusted model, although the trend towards increased VE after adjusting for health-seeking behaviour remained (Figure 3 and Supplementary Table 5).

**Figure 3.**
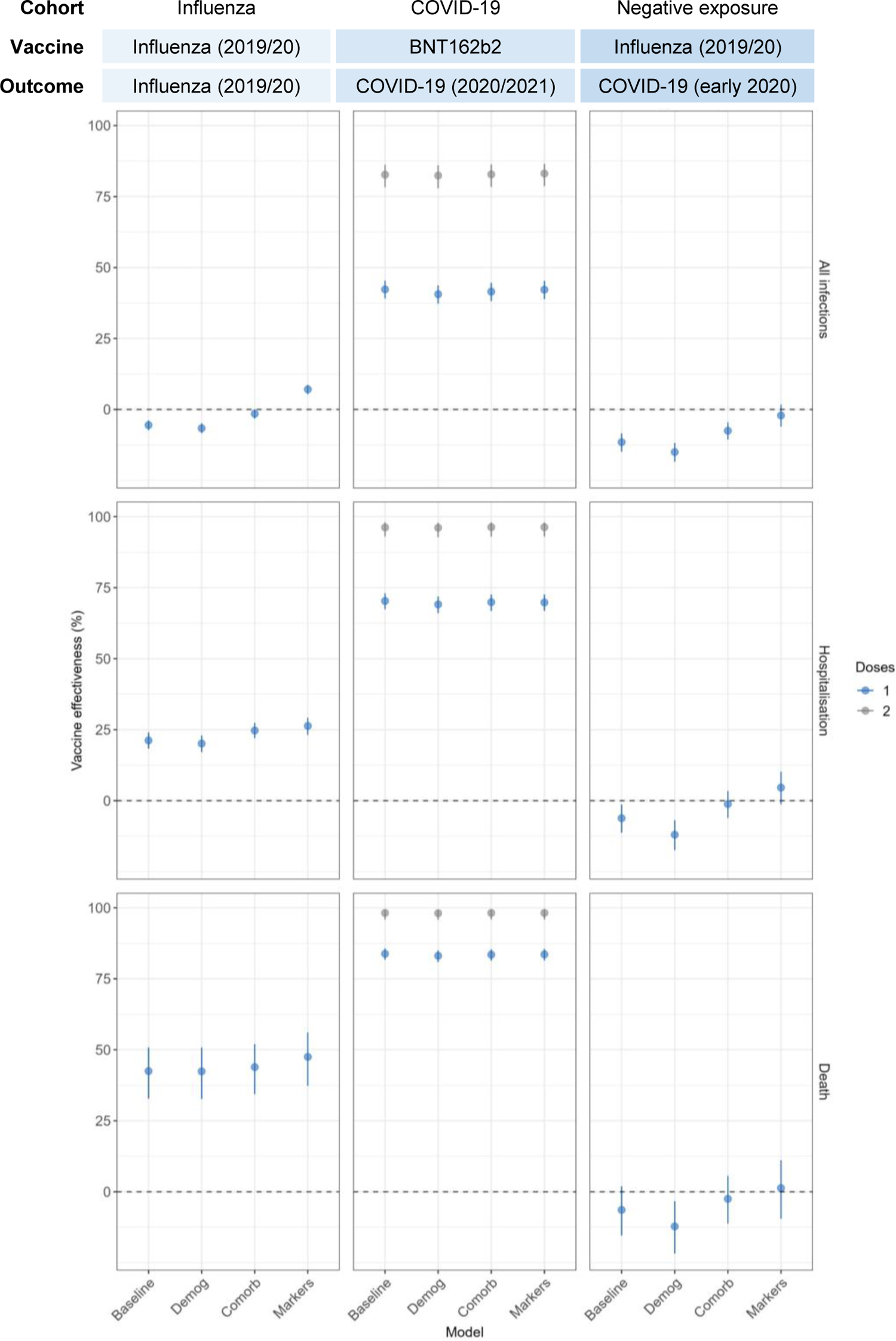
Estimated vaccine effectiveness following sequential confounder adjustment in each study analysis (columns) for each outcome of interest (rows). COVID-19 estimates are only for BNT162b2 versus unvaccinated as ChAdOx1 follow-up data after 2 doses was limited. Baseline models adjusted for polynomial age, sex, region and recent infection. Demography model further adjusted for ethnicity and IMD. Comorbidity models further adjusted for immunosuppressed status and other comorbidities. Marker models further adjusted for markers of health-seeking behaviour and healthcare access. Abbreviations: Comorb: comorbidities; demog: demography.

For the COVID-19 analysis, median (IQR) follow-up time was 113 (0) days. Follow-up time on after first BNT162b2 vaccination was a median of 64 (19) days. Unadjusted event rates ranged from 0.54 per 1,000 person-years after two doses of BNT162b2 vaccination against COVID-19-related death to 95.39 per 1,000 person-years during unvaccinated time against COVID-19 infections (Supplementary Table 4). There was very minimal change in VE from the baseline model to the fully adjusted model that included all 14 markers of health-seeking behaviour and healthcare access (e.g. 2-dose VE of 82.7% [95%CI: 78.3,86.2] and 83.1% [95%CI: 78.7,86.5], respectively). This was also the case for more severe outcomes (e.g. 2-dose VE against hospitalisation of 96.2% [95%CI: 93.0,98.0] and 92.3% [95%CI: 93.0,98.0] for baseline and fully adjusted models, respectively). For ChAdOx1, there was very limited follow-up time after two doses; however, we observed consistent 1-dose VE estimates across baseline and incrementally adjusted models (Supplementary Table 5).

For the negative exposure analysis, median (IQR) follow-up time was 341 (0) days. For those with a history of influenza vaccination, follow-up time was also 341 (0) days. Unadjusted event rates ranged from 1.39 per 1,000 person-years for unvaccinated individuals against COVID-19-related death to 13.77 per 1,0000 person-years for influenza vaccinated individuals against COVID-19-related infections (Supplementary Table 4). We observed a negative VE for the effect of influenza vaccinations against COVID-19 for all baseline and demography-adjusted models (e.g. −6.4% [95%CI: −15.4,-1.9] and −12% [95%CI: −17.4,-6.9], respectively, for VE against COVID-19-related mortality). For infections, negative VE persisted after adjusting for comorbidities (−7.5% [95%CI: −10.6,-4.5]), but not after adjusting for health-seeking behaviour and healthcare access (−2.1% [95%CI: −6.0,1.7]). For more severe endpoints, adjusting for comorbidities led to VE estimates consistent with a null finding, and this was also the case after additional adjustment for health-seeking behaviour and healthcare access (Figure 3 and Supplementary Table 5). The sensitivity analyses that additionally included interaction terms between age and age-varying markers did not substantively change VE estimates for any of the analyses (Supplementary Table 5).

## Discussion

### Statement of principal findings

We found that using a range of markers of health-seeking behaviour we were able to address confounding in VE studies of influenza and COVID-19, with consistent findings using a negative exposure. This was assessed using a large cohort of individuals aged ≥66 years in England and confounding from health-seeking behaviour and healthcare access was adjusted for using proxy markers identified in UK EHRs. We found that influenza and COVID-19 vaccination uptake was higher in those with active health-seeking behaviour and better access to healthcare services. For vaccine effectiveness the impact of health-seeking behaviour and healthcare access varied by context. For influenza in a pre-COVID-19 pandemic context (influenza VE during the 2019/2020 season), we found that VE against infections (including those identified in primary care settings was underestimated when health-seeking behaviour and healthcare access was not adjusted for. This confounding was less apparent for more severe disease endpoints. For COVID-19 in a pandemic context (during the early stages of COVID-19 vaccine implementation), we found that minimally adjusted models were very similar to more complex models, and additional adjustment for health-seeking behaviour and healthcare access did not substantively change effect estimates. Residual confounding was initially present and successfully removed by adjusting for health-seeking behaviour in a negative control analysis exploring the VE of pre-pandemic influenza vaccination against SARS-CoV-2 infections early in the pandemic.

### Strengths and weaknesses of the study

We used a large cohort of individuals and conducted a harmonised analysis with consistent variable definitions and modelling approaches in both pre- and during the COVID-19 pandemic. Although previous VE analyses have adjusted for variables that aim to capture health-seeking behaviour (e.g. GP visits or SARS-CoV-2 testing behaviour),(7, 8, 11) we included a set of proxy markers that were based on a theoretical model published previously(12), which provided a more systematic approach to adjusting for this complex phenomenon. These markers are captured in routinely collected data sources and therefore can be used in other datasets and in other study designs where health-seeking behaviour is a concern. We were also able to identify and quantify residual confounding using a cohort design and negative exposure and demonstrate the impact of adjusting for health-seeking behaviour and healthcare access to address this. We also used nested endpoints which allowed us to assess the implications of confounding on varying outcome severities.

Despite these strengths, we assessed health-seeking behaviour and healthcare access at index date, but this might change over time, especially for the COVID-19 analysis, in which risk perception likely influenced health behaviours. There could potentially be other time-varying confounding(29) if for example, non-vaccination leads to infection and therefore temporary ineligibility for vaccination(30). In addition, there may be some selection bias in the negative exposure study as history of influenza vaccination was identified pre-1 September 2019, but COVID-19 outcomes were identified after this date. This could have potentially led to a slight underestimate of COVID-19 infections in those without influenza vaccination.

### Strengths and weaknesses in relation to other studies, discussing important differences in results

VE estimates from the comorbidity adjusted models were similar to previous estimates from other observational studies during the same time period. For influenza, previous test-negative case control studies estimated VE against virology confirmed disease in the 2019/2020 season to be 22.7% (95%CI: −38.5,56.9%),(31) which is broadly consistent with our estimated VE (adjusted for socio-demographic variables and comorbidities i.e., the most comparable) against ARI/ILI hospital/death 24.7% (95%CI: 22.0 - 27.4). For COVID-19, Whitaker et al.(11) conducted both a cohort and test-negative study to estimate effectiveness of COVID-19 vaccinations against polymerase chain reaction confirmed COVID-19 from 8 December 2020 until 16^th^ May 2021. They estimated VE amongst individuals aged ≥65 years after 2 doses of BNT162b2 to be 84.7% (95%CI: 77.7%,89.5) for the cohort and 79.8% (95%CI: 69.0,86.8) in the test-negative design– consistent with our all infection estimate (adjusted for socio-demographic variables and comorbidites) of 82.8% (95%CI: 78.4 - 86.3).

Our results differ to two US Medicare studies that assessed adjusting for proxy markers of health-seeking behaviour and healthcare access on VE estimates(32, 33). Whilst the two US studies both saw a decrease in VE after adjusting for confounding from health-seeking behaviour and healthcare access, we saw an increase in VE for influenza. These differences could be because of the differences in healthcare settings (i.e., nationwide free healthcare versus a premium paid healthcare) or differences in the type of dataset (i.e., EHRs versus insurance claims). It could also be that the US studies used a smaller set of markers than the current study, and therefore it could be that residual confounding remained. One of the US studies(33) used pre-season influenza estimates as a negative control outcome and still found significant residual confounding even after adjustment for health-seeking behaviour and healthcare access as well as other socio-demographic variables and proxies for frailty (32% [95%CI: 30-33%]).

For the negative control analysis we assumed that there was no causal mechanism between influenza vaccination and COVID-19 infection, or that any plausible causal association was minimal in effect. Some studies with non-specific COVID-19 outcomes have shown there to be a minor protective effect of the influenza vaccination against COVID-19 infection. A meta-analysis of 16 observational studies reported pooled odds ratios of influenza vaccination against COVID-19 infection of 0.86 (95%CI: 0.81,0.91)(34). The authors of this study suggested that individuals with influenza vaccinations are likely to have better health-seeking behaviours and healthcare access and may therefore be more compliant with COVID-19 prevention measures. A recent observational study conducted using administrative data in Canada reported reduced COVID-19 infections amongst those with an influenza vaccination, but also reported the same trend for COVID-19 infections amongst those with a previous health examination (adjusted HR:D0.85 [95%CI: 0.78-0.91])(35) and summarised that this provided evidence of confounding from health-seeking behaviour and healthcare access.

Our study showed an opposite trend to the Canadian study for our negative exposure. Prior to adjusting for confounding from our proxies of health-seeking behaviour and healthcare access, our negative exposure analysis showed a negative VE. This likely represents those with active health-seeking behaviour being more likely to present to primary care when they contract COVID-19-like symptoms compared to those with poorer health-seeking behaviours. When we adjusted for markers of health-seeking behaviour and healthcare access, we saw a null effect representing that residual confounding had been removed.

Although prior influenza vaccination may be relevant as a negative exposure for COVID-19 in the first wave of the pandemic, this may become less appropriate over time due to the strong association between influenza and COVID-19 vaccinations.

### Meaning of the study: possible explanations and implications for clinical and policymakers

We hope the findings from this study will be useful to other observational researchers. Authors will be able to characterise the health behaviours of their population, to identify the strength and direction of this confounding from health-seeking behaviour and healthcare access on their effect estimates, and to account for any identified confounding using the markers provided. We believe that particularly for seasonal influenza and COVID-19, these markers could be helpful for future researchers to provide more accurate annual VE and cost-effectiveness estimates. Other areas of research whether we believe these markers might be important are for chronic conditions such as chronic kidney disease and diabetes, for which health-seeking behaviour and healthcare access has previously been shown to influence timeliness of seeking care and self-management of these conditions.(36, 37) Usefulness of these markers is likely to vary by context and study type. For example, they are likely to be more useful for routine rather than pandemic VE estimates. As we saw for the COVID-19 during the pandemic after early vaccine deployment, sequential model adjustments for additional socio-demographic and health-seeking behaviour and healthcare access variables had limited impact on VE estimates. This is likely due to the high-risk perception of the virus and high testing and vaccination capacity during this time which meant that prior health-seeking behaviour and healthcare access was less influential. These markers are likely to be more useful in older cohorts as many of the markers used are relevant to individuals aged ≥65 years. Marker will have to be adapted when including younger individuals.

The descriptive results of this study are likely to be useful to clinicians and policymakers that are interested in identifying characteristics of individuals that are more likely to take up vaccinations and other nationwide programmes. We showed that individuals that take up UK nationwide screening programmes and NHS health checks are more likely to be those that get vaccinated. We also showed that individuals with previous influenza vaccinations are more likely to take up vaccinations in subsequent seasons. Governments could use this information to target groups of individuals to improve health inequality. Policy makers and clinicians can also use the findings from this study to increase their reassurance that during pandemic contexts, minimally adjusted VE estimates from observational studies, which are rapidly produced to guide decision making, are likely to be a close reflection of final VE estimates.

### Unanswered questions and future research

It would also be useful to understand how well these markers perform in different settings, study types and for different research questions, including designs that explicitly account for time-varying confounders(30). The rate of uptake of preventative services, including vaccinations, are likely to vary by country and therefore the usefulness of these markers is likely to differ by settings.

## Conclusion

We have identified markers in UK EHRs that can be used to quantify and adjust for confounding from health-seeking behaviour and healthcare access in observational research. Adjusting for health-seeking behaviour had a limited influence on estimates of COVID-19 VE during the pandemic and early stages of the vaccine roll-out. For seasonal influenza VE, severe outcomes were robust to confounding from health-seeking behaviour, but VE against influenza infections identified in primary care were underestimated prior to adjustment for health-seeking behaviour. Residual confounding was also removed as demonstrated in a negative exposure analysis of history of influenza vaccination against COVID-19 infections (pre COVID-19 vaccine roll out).

## Funding

SG, EPKP, NA, JLW and HIM are funded by the National Institute of Health and Care Research (NIHR) Health Protection Research Unit in Vaccines and Immunisation (grant reference: NIHR200929), a partnership between UK Health Security Agency (UKHSA) and London School of Hygiene and Tropical Medicine. The views expressed are those of the author(s) and not necessarily those of the NIHR, UKHSA or the Department of Health and Social Care.

## Disclaimer

The views expressed are those of the authors and not necessarily those of the NHS, the NIHR, or UKHSA.

## Competing interests

SG is also a part-time salaried employee of Evidera, which is a business unit of Pharmaceutical Product Development (PPD), part of Thermo Fisher Scientific.

## Data Availability

These data were obtained from the Clinical Practice Research Datalink, provided by the UK Medicines and Healthcare products Regulatory Agency. The authors’ licence for using these data does not allow sharing of raw data with third parties. Information about access to Clinical Practice Research Datalink data is available here: https://www.cprd.com/research-applications. Codelists for this study are available at https://doi.org/10.17037/DATA.00003684 and code at https://github.com/grahams99/Health-seeking-behaviour.

## Notes

### Author Declarations

Our study was approved by CPRDs (#21_000737) and LSHTMs (#28169) independent ethics review committees. To meet CPRD patient confidentiality requirements we redacted counts relating to less than 5 individuals and conducted secondary suppression where necessary.

