## Supplementary information for "Quantifying and adjusting for confounding from health-seeking behaviour and healthcare access in observational research"

### Tables

**Supplementary Table 1. Overview of study design and population selection in all analyses**

|  | **COVID-19** | **Influenza** | **Negative exposure** |
| --- | --- | --- | --- |
| **Index date/start of follow-up for all individuals** | 8 December 2020 | 1 September 2019 | 1 January 2020 |
| **End of follow-up** | Earliest of death, transfer out of the practice, end of data availability (29 March 2021), date of first COVID-19 vaccination that was neither BNT162b2 or ChAdOx1 or date of second heterologous vaccination (Figure 1). | Earliest of death, transfer out of the practice or end of influenza season (defined as 29 February 2020; Figure 1). | Earliest of death, transfer out of the practice or start of COVID-19 vaccination availability (7 December 2020; Figure 1). |
| **Population selection** | **Inclusion criteria:**   1. Aged ≥66 years on 1 September 2019. 2. Registration start date one year before index date. 3. Acceptable flag. 4. Eligible for HES APC and ONS linkage.   **Exclusion criteria:**   1. Registration end date or death before index date. 2. Indeterminate sex. | | |
| **Additional criteria** | **Exclusion criteria:**   1. Individuals with a COVID-19 vaccination that occurred before 8 December 2020 (as these were likely to be trial participants with different risk). |  |  |
| **Outcomes** | - SARS-CoV-2 infection (GP visit, hospital visit or death with COVID-19 specific medcode or ICD-19 code). - Hospital visit or death with COVID-19 specific ICD-10 code. - Death with COVID-19 specific ICD-10 code.   Both suspected and confirmed COVID-19 medcodes were used since we wanted a consistent definition with Negative Exposure (see Negative Exposure column). | - Acute respiratory infection or influenza/influenza-like-illness infection (ARI/ILI; GP visit, hospital visit or death with ARI/ILI specific medcode or ICD-19 code). - Hospital visit or death with ARI/ILI specific ICD-10 code. - Death with ARI/ILI specific ICD-10 code. | - SARS-CoV-2 infection (GP visit, hospital visit or death with COVID-19 medcode or ICD-10 specific code). - Hospital visit or death with COVID-19 specific ICD-10 code. - Death with COVID-19 specific ICD-10 code.   Both suspected and confirmed COVID-19 medcodes were used since majority of the outcome period occurred before the availability of widespread free polymerase chain reaction testing in the UK. |
| **Exposures** | - 1 or 2 doses of BNT162b2 or ChAdOx1 from 8 December 2020 onwards. We only used prodcodes to identify these, since it was not possible to identify the brand using medcodes. | - 1 dose of any influenza vaccination in the 2019/2020 influenza season (1 September 2019-29 February 2020). Both prodcodes and medcodes were used to identify influenza vaccinations. An algorithm was developed for code that occurred on the same day – see Supplementary Table 2. | - A history of 1 dose of any timely influenza vaccination in the 2019/2020 season (1 September 2019-31 December 2019; binary, assessed at baseline). Both prodcodes and medcodes were used to identify influenza vaccinations. An algorithm was developed for code that occurred on the same day – see Supplementary Table 2. |
| **Variables described at index date** | - Age in years (categorised as 65-69, 70-74, 75-79, 80-84, 85-89, 90-95, 95+) - Sex (male, female) - Recent infection (COVID-19 VE analysis: covid-19 infection in the last 3-months; Influenza VE analysis: influenza infection in the previous influenza season [1 April 2018 to 31 August 2019]) - IMD (categorised as from 1 [least deprived] to 5 [most deprived]) - Ethnicity (categorised as Asian, Black, Missing, Mixed, Other and White) - ‘Influenza at risk’ conditions (immunosuppressed status and other – see below).   Comorbidities in influenza ‘at-risk’ groups were identified according to Greenbook chapter 19(26) in the pre-index period using medcodes (unless otherwise specified) and the below specified lookback periods:  Immunosuppressed status:   - Organ recipient: any time prior to index. - Immunosuppression therapies: biologic within the one year prior to index; or corticosteroids >40mg prednisolone per day for more than 1 week or corticosteroids >20mg prednisolone per day for more than 14 day or methotrexate >25mg per week; azathioprine >3.0mg/kg/day; 6-mercaptopurine >1.5mg/kg/day or corticosteroid injections; other disease-modifying antirheumatic drugs or other immunosuppressant medications in the 3-months prior to index. - Other immunosuppression: any time prior to index.   Other conditions:   - Chronic liver disease: any time prior to index. - Chronic cardiac disease: any time prior to index. - Chronic respiratory disease: any time prior to index. - Asthma: 3-months prior to index. - Diabetes mellitus: any time prior to index. - Chronic kidney disease: dialysis or transplant any time prior to index; or latest chronic kidney disease code is stage 3-5 (and not 1-2); or latest serum creatinine test result value ≤60 mL/min/1.73m2. - Chronic neurological disease: any time prior to index. - Severe obesity: latest body mass index recording prior to index ≥40 kg/m^2^. - Severe mental conditions: any time prior to index. - Severe learning disability: any time prior to index.   Code lists from Davidson et al, 2021(23) were utilised. The code lists from can be found listed on London School of Hygiene and Tropical Medicine (LSHTM) data compass: <https://datacompass.lshtm.ac.uk/id/eprint/2240/>. Previous estimates have shown that use of medcodes alone give plausible prevalence estimates.(38) | | |
| **Markers of health-seeking behaviour and healthcare access and look back period** | Markers were previously identified in Graham et al.(12)  All of these conditions were identified using medcodes, ICD-10, OPCS or prodcodes identified in CPRD Aurum or HES APC. The same operational definitions from Graham et al.(12) were utilised:  :   - Abdominal aortic aneurysm screening (sex specific): any time prior to index date. - Breast cancer screening (sex specific): the last 4 years that they were age-eligible for screening prior to index date. - Cervical cancer screening (sex specific): the last 6 years that they were age-eligible for screening prior to index date. - Bowel cancer screening: from the last 3 years that they were age-eligible for screening prior to index date. - NHS healthcare checks: the last 6 years that they were age-eligible for NHS health checks prior to index date. - Influenza vaccination: from 1 September 2018-31 March 2019 (influenza and negative exposure analysis); from 1 September 2019-31 March 2020 (COVID-19 analysis). - Pneumococcal vaccination: any time prior to index. - Prostate specific antigen testing: the last three years prior to index. - Bone density scans: the last three years prior to index. - GP practice visits: the last year prior to index. - Did not attend primary care visit: the last year prior to index. - Low value procedures: the last year prior to index. - Hospital visit for ambulatory care sensitive conditions: the last five years prior to index. - Blood pressure measurements: the last year prior to index.   Code lists from Graham et al.(12) were utilised. The code lists from this project can be found listed on LSHTM data compass: <https://doi.org/10.17037/DATA.00003684>. | | |

Abbreviations: HES: Hospital Episode Statistics; ICD-10: International Classification of Diseases 10^th^ revision; IMD: index of multiple deprivation; LSHTM: London School of Hygiene and Tropical Medicine; OPCS: Operating Procedure Codes Supplement.

**Supplementary Table 2. Influenza vaccination algorithm**

| **Combination of codes on same day** | **Total number of vaccination events** | **Decision** | **Rationale** |
| --- | --- | --- | --- |
| Given and neutral | 725432 | Record as valid vaccination. |  |
| Given and absent | 1149 (of these only 924 are the first vaccination dose) | Do not record as valid vaccination. | The prevalence of this marker may be underestimated very slightly, however, in this instance better to be more specific than sensitive when it comes to confounders(39). |
| Given and adverse | 14 | Record as valid vaccination. | Likely that this patient received the vaccination, but then had an adverse event on the same day. |
| Given and product | 395881 | Record as valid vaccination. |  |
| Given and given with lag | 4700 | Record as valid vaccination. |  |
| Neutral and absent | 1178 | Do not record as valid vaccination. |  |
| Neutral and adverse | 9 | Record as valid vaccination. |  |
| Neutral and product | 295746 | Record as valid vaccination. |  |
| Neutral and given lag | 7748 | Record as valid vaccination. |  |
| Absent and adverse | 27 | Do not record as valid vaccination. | Likely that these patients are reporting a previous adverse event as the reason for not wanting to get vaccinated. |
| Absent and product | 218 (of these only 194 are first vaccination dose) | Do not record as valid vaccination. | The prevalence of this marker will be underestimated very slightly, however, in this instance better to be more specific than sensitive when it comes to confounders(39). |
| Absent and given with lag | 570 | Record as valid vaccination. | This most likely reflects that the reason a vaccine wasn’t given on the event date is that the patient had already had it elsewhere. We can be reasonably confident that the patient was vaccinated, but we don’t know the exact date. |
| Adverse and product | <5 | Record as valid vaccination. | Likely that these patients received the vaccination, but then had an adverse event on the same day. |
| Adverse and given with lag | 0 | Ignore – no events. |  |
| Given with lag and product | 784 | Record as valid vaccination. |  |

Note: this table is attributed from Graham et al.(12) Since influenza vaccinations can be identified using both medcodes and prodcodes and since medcodes do not always insinuate presence of a vaccination, an algorithm was developed for combinations of codes that occurred on the same day. Medcodes were separated into those that were clearly given (“given” or “administered”), given with delay (“given” or “administered” but evidence this occurred in another setting previously), neutral (vaccination mentioned but no “given” or “administered”) and absent (vaccination “refused” or “not consented). Then we looked at vaccination events (using both medcodes and prodcodes) that were recorded on the same date and categorised these according to the above framework. We found 33.5% of individuals with >1 influenza prodcode or medcode during the 2019/2020 season. However, since individuals can have both a prodcode and medcode recorded for each vaccination event these were unlikely to be true vaccination events and therefore were ignored. Cells with <5 individuals are redacted due to CPRD’s patient confidentiality requirements and secondary suppression has occurred where necessary.

**Supplementary Table 3. Baseline characteristics stratified in overall analysis populations.**

| **Variable** | **Category** | **COVID-19 analysis population**  **N=1,796,667** | **Influenza analysis population**  **N=1,991,284** | **Negative exposure analysis population**  **N=1,946,943** |
| --- | --- | --- | --- | --- |
| **Age category in years, N (%)** | 65-69 | 448,063 (22.5%) | 319,958 (17.8%) | 442,923 (22.7%) |
|  | 70-74 | 557,599 (28.0%) | 540,955 (30.1%) | 550,104 (28.3%) |
|  | 75-79 | 401,290 (20.2%) | 391,299 (21.8%) | 394,068 (20.2%) |
|  | 80-84 | 295,492 (14.8%) | 278,776 (15.5%) | 287,737 (14.8%) |
|  | 85-89 | 181,835 (9.1%) | 169,263 (9.4%) | 173,832 (8.9%) |
|  | 90-95 | 80,943 (4.1%) | 74,430 (4.1%) | 75,278 (3.9%) |
|  | 95+ | 26,062 (1.3%) | 21,986 (1.2%) | 23,001 (1.2%) |
| **Sex, N (%)** | Female | 1,075,723 (54.0%) | 973,794 (54.2%) | 1,052,528 (54.1%) |
|  | Male | 915,561 (46.0%) | 822,873 (45.8%) | 894,415 (45.9%) |
| **Ethnicity, N (%)** | Asian | 67,961 (3.4%) | 63,829 (3.6%) | 67,270 (3.5%) |
|  | Black | 36,912 (1.9%) | 33,981 (1.9%) | 36,454 (1.9%) |
|  | Missing | 98,874 (5.0%) | 89,889 (5.0%) | 97,103 (5.0%) |
|  | Mixed | 10,072 (0.5%) | 9,209 (0.5%) | 9,948 (0.5%) |
|  | Other | 17,711 (0.9%) | 16,533 (0.9%) | 17,593 (0.9%) |
|  | White | 1,759,754 (88.4%) | 1,583,226 (88.1%) | 1,718,575 (88.3%) |
| **Region, N (%)** | East Midlands | 95,413 (4.8%) | 35,304 (2.0%) | 38,029 (2.0%) |
|  | East of England | 263,825 (13.2%) | 81,497 (4.5%) | 88,480 (4.5%) |
|  | London | 67,278 (3.4%) | 239,370 (13.3%) | 260,228 (13.4%) |
|  | North East | 381,592 (19.2%) | 62,453 (3.5%) | 66,590 (3.4%) |
|  | North West | 438,407 (22.0%) | 347,533 (19.3%) | 376,009 (19.3%) |
|  | South East | 270,484 (13.6%) | 395,424 (22.0%) | 432,893 (22.2%) |
|  | South West | 357,363 (17.9%) | 245,084 (13.6%) | 259,419 (13.3%) |
|  | West Midlands | 74,805 (3.8%) | 323,547 (18.0%) | 351,397 (18.0%) |
|  | Yorkshire and The Humber | 11 (0.0%) | 66,352 (3.7%) | 73,860 (3.8%) |
|  | Unknown | 499,873 (25.1%) | 103 (0.0%) | 38 (0.0%) |
| **IMD, N (%)** | 1 (least deprived) | 467,781 (23.5%) | 458,067 (25.5%) | 492,109 (25.3%) |
|  | 2 | 399,311 (20.1%) | 420,214 (23.4%) | 457,789 (23.5%) |
|  | 3 | 344,964 (17.3%) | 357,465 (19.9%) | 386,545 (19.9%) |
|  | 4 | 279,355 (14.0%) | 310,988 (17.3%) | 336,847 (17.3%) |
|  | 5 (most deprived) | 95,413 (4.8%) | 249,933 (13.9%) | 273,653 (14.1%) |
| **Influenza ‘at-risk’ conditions, N (%)** | Immunosuppressed status | 55,153 (2.8%) | 44,055 (2.5%) | 62,795 (3.2%) |
|  | Other comorbidities* | 1,103,284 (55.4%) | 1,004,589 (55.9%) | 1,084,662 (55.7%) |
| **Markers of health-seeking behaviour and healthcare access, N (%)** | AAA screen | 231,088 (11.6%) | 214,942 (12.0%) | 227,316 (11.7%) |
|  | Breast screen | 346,116 (17.4%) | 327,105 (18.2%) | 343,720 (17.7%) |
|  | Cervical screen | 397,303 (20.0%) | 362,943 (20.2%) | 388,994 (20.0%) |
|  | Bowel screen | 1,439,412 (72.3%) | 1,354,825 (75.4%) | 1,424,238 (73.2%) |
|  | NHS health checks | 372,244 (18.7%) | 321,029 (17.9%) | 363,677 (18.7%) |
|  | Influenza vaccine† | 1,460,391 (73.3%) | 1,363,429 (75.9%) | 1,427,057 (73.3%) |
|  | Pneumococcal vaccine | 1,242,359 (62.4%) | 1,158,676 (64.5%) | 1,233,497 (63.4%) |
|  | PSA test | 352,272 (17.7%) | 366,640 (20.4%) | 363,037 (18.6%) |
|  | Bone density scan | 100,892 (5.1%) | 115,237 (6.4%) | 108,540 (5.6%) |
|  | Low value procedures | 358,881 (18.0%) | 537,129 (29.9%) | 435,940 (22.4%) |
|  | Primary care DNA | 601,896 (30.2%) | 935,986 (52.1%) | 731,921 (37.6%) |
|  | Hospital visit for ambulatory care sensitive conditions | 190,136 (9.5%) | 190,805 (10.6%) | 195,863 (10.1%) |
|  | Blood pressure test | 1,470,006 (73.8%) | 1,526,971 (85.0%) | 1,563,575 (80.3%) |
|  | GP visits | 1,844,823 (92.6%) | 1,741,469 (96.9%) | 1,847,410 (94.9%) |

Abbreviations: AAA: abdominal aortic aneurysm; DNA: did not attend; GP: general practice; IMD: index of multiple deprivation; N: numerator; NHS: National Health Service; PSA: prostate specific antigen; VE: vaccine effectiveness.

*Other comorbidities includes: chronic liver disease, chronic cardiac disease, chronic respiratory disease, asthma, diabetes mellitus, chronic neurological disease, chronic kidney disease, severe obesity, severe mental conditions and severe learning disability. For more information on how these were defined see **Supplementary Table 1.**

†Influenza vaccination that occurred in the influenza season prior to index date. For COVID-19 this was an influenza vaccination that occurred 1 September 2019 – 31 March 2020; for Influenza and Negative Exposure this was an influenza vaccination that occurred 1 September 2018-31 March 2019.

Notes: we are comparing individuals with ≥1 vaccination versus no vaccination throughout whole of follow-up. Cells with <5 individuals are redacted due to CPRD’s patient confidentiality requirements and secondary suppression has occurred where necessary.

**Supplementary Table 4. Unadjusted rates for each analysis and outcome**

|  | **Infections** | | | **Hospital or death** | | | **Death** | | |
| --- | --- | --- | --- | --- | --- | --- | --- | --- | --- |
| **Exposure** | **Events** | **Person years** | **Rate per 1,000 person years** | **Events** | **Person years** | **Rate per 1,000 person years** | **Events** | **Person years** | **Rate per 1,000 person years** |
| **COVID-19 cohort analysis** | | | | | | | | | |
| **BNT162b2** | | | | | | | | | |
| Unvaccinated | 14,516 | 152,174.8 | 95.39 | 5525 | 153,293.7 | 36.04 | 3076 | 153,698.3 | 20.01 |
| One dose | 2,381 | 107,497.5 | 22.15 | 622 | 10,8495 | 5.73 | 366 | 10,8655 | 3.37 |
| Two doses | 77 | 11,062.77 | 6.96 | 10 | 11,121.57 | 0.9 | 6 | 11,128.44 | 0.54 |
| **ChAdOx1** | | | | | | | | | |
| Unvaccinated | 22559 | 186384.6 | 121.03 | 7206 | 188467.5 | 38.23 | 3097 | 189131.7 | 16.37 |
| One dose | 1626 | 90524.3 | 17.96 | 459 | 91875.61 | 5 | 311 | 92119.05 | 3.38 |
| Two doses | [Redacted] | 76.75 | [Redacted] | 0 | 79.27 | 0 | 0 | 79.5 | 0 |
| **Influenza cohort analysis** | | | | | | | | | |
| Unvaccinated | 40420 | 427719.1 | 94.5 | 7451 | 432900.5 | 17.21 | 364 | 434087.6 | 0.84 |
| One dose | 54210 | 462753.6 | 117.15 | 9263 | 476088.3 | 19.46 | 428 | 478248.4 | 0.89 |
| **Negative exposure cohort analysis** | | | | | | | | | |
| Unvaccinated | 6192 | 528783.4 | 11.71 | 2257 | 528788.2 | 4.27 | 736 | 528789.8 | 1.39 |
| One dose | 22095 | 1604967 | 13.77 | 8176 | 1604968 | 5.09 | 2865 | 1604973 | 1.79 |

Notes: events represents the total number of events identified in the study follow up period. Population represents the total number of individuals included in each group. Person years is the total time in years until end of follow-up. It should be noted that for the COVID-19 and influenza analyses person years is time whilst unexposed/exposed, whereas for the negative exposure analysis, since we used a binary exposure at baseline, person-years is all available follow-up from index until end of follow-up. Rate is calculated as the total number of events divided by the total person years multiplied by 1,000. Cells with <5 events are redacted due to CPRD’s patient confidentiality requirements and secondary suppression has occurred where necessary.

**Supplementary Table 5. Vaccine effectiveness estimates**

| **Model** | **All infections**  **VE (95%CI)** | **Hospitalisation or death**  **VE (95%CI)** | **Death**  **VE (95%CI)** |
| --- | --- | --- | --- |
| **Influenza** | | | |
| Baseline | -5.5 (-7.2, -3.9) | 21.2 (18.3, 24.0) | 42.5 (32.8, 50.8) |
| Demography | -6.6 (-8.3, -4.9) | 20.1 (17.1, 22.9) | 42.4 (32.7, 50.8) |
| Comorbidities | -1.5 (-3.2, 0.1) | 24.7 (22.0, 27.4) | 43.9 (34.4, 52.0) |
| Markers | 7.1 (5.4, 8.7) | 26.3 (23.1, 29.2) | 47.5 (37.3, 56.1) |
| Sensitivity | 7.2 (5.5, 8.9) | 26.4 (23.3, 29.4) | 47.4 (37.1, 55.9) |
| **COVID-19 (BNT162b2) dose one** | | | |
| Baseline | 42.3 (39.1, 45.4) | 70.3 (67.4, 73.0) | 83.8 (81.7, 85.7) |
| Demography | 40.6 (37.3, 43.7) | 69.1 (66, 71.9) | 83.1 (80.9, 85.0) |
| Comorbidities | 41.5 (38.2, 44.6) | 69.9 (66.8 - 72.6) | 83.5 (81.4, 85.4) |
| Markers | 42.2 (38.9, 45.3) | 69.8 (66.8, 72.6) | 83.6 (81.5, 85.5) |
| Sensitivity | 42.1 (38.9, 45.2) | 69.8 (66.8, 72.5) | 83.6 (81.5, 85.5) |
| **COVID-19 (BNT162b2) dose two** | | | |
| Baseline | 82.7 (78.3, 86.2) | 96.2 (93.0, 98.0) | 98.2 (95.9, 99.2) |
| Demography | 82.4 (77.9, 86.0) | 96.1 (92.8, 97.9) | 98.1 (95.8, 99.2) |
| Comorbidities | 82.8 (78.4, 86.3) | 96.3 (93.0, 98.0) | 98.2 (95.9, 99.2) |
| Markers | 83.1 (78.7, 86.5) | 96.3 (93.0, 98.0) | 98.2 (95.9, 99.2) |
| Sensitivity | 83.0 (78.7, 86.5) | 96.2 (93.0, 98.0) | 98.2 (95.9, 99.2) |
| **COVID-19 (ChAdOx1)** | | | |
| Baseline | 7.6 (0.4, 14.2) | 24.9 (14.7 - 33.9) | 51.0 (43.0 - 57.8) |
| Demography | 5.0 (-2.3 - 11.9) | 21.9 (11.3 - 31.2) | 49.3 (41.1 - 56.4) |
| Comorbidities | 6.5 (-0.8 - 13.2) | 23.5 (13.1 - 32.7) | 50.4 (42.3 - 57.3) |
| Markers | 9.6 (2.6 - 16.2) | 25.7 (15.6 - 34.6) | 52.5 (44.8 - 59.2) |
| Sensitivity | 8.8 (1.7 - 15.4) | 25.3 (15.2 - 34.3) | 52.4 (44.6 - 59.1) |
| **Negative Exposure** | | | |
| Baseline | -11.5 (-14.8 - -8.4) | -6.2 (-11.3 - -1.4) | -6.4 (-15.4 - 1.9) |
| Demography | -15 (-18.4 - -11.8) | -12 (-17.4 - -6.9) | -12.2 (-21.7 - -3.3) |
| Comorbidities | -7.5 (-10.6 - -4.5) | -1.2 (-6.1 - 3.4) | -2.5 (-11.2 - 5.6) |
| Markers | -2.1 (-6.0 - 1.7) | 4.6 (-1.3 - 10.2) | 1.3 (-9.5 - 11.1) |
| Sensitivity | -2.2 (-6.1 - 1.5) | 4.5 (-1.5 - 10.1) | 1.2 (-9.6 – 11.0) |

Notes: baseline: adjusted for polynomial age, sex, region and recent infection. Demography model: baseline model + adjusted for ethnicity and IMD. Comorbidities: demography model + adjusted for immunosuppressed status and other comorbidities. Markers: comorbidities model + adjusted for each marker of health-seeking behaviour and healthcare access separately with sex interactions for sex-specific markers. Sensitivity: markers model + age interactions for AAA screening, bowel cancer screening, NHS health checks and ACS conditions. Vaccine effectiveness is estimated as (1-HR)*100.

Abbreviations: SES: socioeconomic status.

**Supplementary Table 8. Amongst vaccinated individuals only, median days from index to influenza vaccination stratified by marker status and age category**

|  | **Presence of marker** | | **Absence of marker** | |
| --- | --- | --- | --- | --- |
| **Marker** | **Age category** | **Median (q1 – q3)** | **Age category** | **Median (q1 – q3)** |
| AAA Screen Males | Overall | 50 (39 - 67) | Overall | 50 (38 - 66) |
|  | 65-69 | 52 (39 - 69) | 65-69 | 53 (40 - 72) |
|  | 70-74 | 50 (39 - 67) | 70-74 | 51 (39 - 67) |
|  | 75-79 | 48 (38 - 62) | 75-79 | 49 (38 - 65) |
|  | 80-84 | 48 (37 - 62) | 80-84 | 48 (38 - 64) |
|  | 85+ | 47 (37 - 62) | 85+ | 50 (39 - 67) |
| Bowel Cancer Screen | Overall | 50 (39 - 67) | Overall | 51 (39 - 68) |
|  | 65-69 | 52 (39 - 69) | 65-69 | 53 (41 - 72) |
|  | 70-74 | 50 (39 - 67) | 70-74 | 51 (40 - 68) |
|  | 75-79 | 48 (38 - 65) | 75-79 | 51 (39 - 66) |
|  | 80-84 | 48 (38 - 64) | 80-84 | 50 (38 - 66) |
|  | 85+ | 50 (38 - 65) | 85+ | 52 (39 - 69) |
| Breast Cancer Screen Females | Overall | 51 (39 - 67) | Overall | 51 (39 - 68) |
|  | 65-69 | 51 (39 - 68) | 65-69 | 52 (39 - 69) |
|  | 70-74 | 50 (38 - 65) | 70-74 | 50 (39 - 67) |
|  | 75-79 | 48 (38 - 64) | 75-79 | 50 (39 - 66) |
|  | 80-84 | 50 (38 - 65) | 80-84 | 50 (39 - 66) |
|  | 85+ | 50 (39 - 66) | 85+ | 52 (40 - 71) |
| Cervical Cancer Screen Females | Overall | 51 (39 - 67) | Overall | 51 (39 - 67) |
|  | 65-69 | 52 (40 - 69) | 65-69 | 51 (39 - 69) |
|  | 70-74 | 50 (39 - 66) | 70-74 | 50 (39 - 66) |
|  | 75-79 | 50 (38 - 65) | 75-79 | 50 (38 - 65) |
|  | 80-84 | 50 (38 - 65) | 80-84 | 50 (39 - 66) |
|  | 85+ | 51 (39 - 68) | 85+ | 53 (40 - 72) |
| NHS Health Checks | Overall | 50 (39 - 67) | Overall | 51 (39 - 67) |
|  | 65-69 | 52 (39 - 69) | 65-69 | 52 (39 - 69) |
|  | 70-74 | 50 (39 - 66) | 70-74 | 50 (39 - 67) |
|  | 75-79 | 48 (38 - 64) | 75-79 | 50 (38 - 65) |
|  | 80-84 | 48 (38 - 64) | 80-84 | 50 (38 - 65) |
|  | 85+ | 52 (40 - 71) | 85+ | 51 (39 - 68) |
| Influenza Vaccination | Overall | 50 (39 - 67) | Overall | 59 (41 - 86) |
|  | 65-69 | 51 (39 - 67) | 65-69 | 61 (43 - 89) |
|  | 70-74 | 50 (38 - 65) | 70-74 | 59 (41 - 87) |
|  | 75-79 | 48 (38 - 64) | 75-79 | 57 (40 - 83) |
|  | 80-84 | 48 (38 - 64) | 80-84 | 57 (40 - 81) |
|  | 85+ | 51 (39 - 67) | 85+ | 59 (41 - 86) |
| Pneumococcal Vaccination | Overall | 50 (39 - 67) | Overall | 53 (40 - 72) |
|  | 65-69 | 50 (38 - 66) | 65-69 | 54 (41 - 74) |
|  | 70-74 | 48 (38 - 65) | 70-74 | 53 (40 - 72) |
|  | 75-79 | 48 (38 - 64) | 75-79 | 52 (39 - 69) |
|  | 80-84 | 48 (38 - 64) | 80-84 | 52 (39 - 68) |
|  | 85+ | 51 (39 - 68) | 85+ | 53 (40 - 73) |
| ACS Hospital Visit | Overall | 50 (39 - 67) | Overall | 50 (39 - 67) |
|  | 65-69 | 52 (39 - 71) | 65-69 | 52 (39 - 69) |
|  | 70-74 | 51 (39 - 68) | 70-74 | 50 (39 - 67) |
|  | 75-79 | 50 (38 - 67) | 75-79 | 48 (38 - 65) |
|  | 80-84 | 51 (39 - 68) | 80-84 | 48 (38 - 65) |
|  | 85+ | 53 (40 - 73) | 85+ | 51 (39 - 68) |
| Blood Pressure Test | Overall | 50 (39 - 67) | Overall | 52 (39 - 69) |
|  | 65-69 | 51 (39 - 69) | 65-69 | 53 (40 - 72) |
|  | 70-74 | 50 (39 - 66) | 70-74 | 51 (39 - 68) |
|  | 75-79 | 48 (38 - 65) | 75-79 | 51 (39 - 66) |
|  | 80-84 | 48 (38 - 65) | 80-84 | 51 (39 - 67) |
|  | 85+ | 51 (39 - 68) | 85+ | 52 (40 - 71) |
| Bone Density Scan | Overall | 50 (39 - 67) | Overall | 51 (39 - 67) |
|  | 65-69 | 51 (39 - 68) | 65-69 | 52 (39 - 69) |
|  | 70-74 | 50 (38 - 65) | 70-74 | 50 (39 - 67) |
|  | 75-79 | 48 (38 - 64) | 75-79 | 50 (38 - 65) |
|  | 80-84 | 48 (38 - 64) | 80-84 | 50 (38 - 65) |
|  | 85+ | 50 (38 - 66.75) | 85+ | 52 (39 - 69) |
| DNA Primary Care Visit | Overall | 50 (39 - 67) | Overall | 50 (39 - 66) |
|  | 65-69 | 52 (39 - 71) | 65-69 | 52 (39 - 69) |
|  | 70-74 | 51 (38 - 68) | 70-74 | 50 (39 - 66) |
|  | 75-79 | 50 (38 - 66) | 75-79 | 48 (38 - 65) |
|  | 80-84 | 50 (38 - 66) | 80-84 | 48 (38 - 64) |
|  | 85+ | 52 (39 - 70) | 85+ | 51 (39 - 68) |
| GP Visit | Overall | 50 (39 - 67) | Overall | 53 (39 - 72) |
|  | 65-69 | 52 (39 - 69) | 65-69 | 54 (39 - 75) |
|  | 70-74 | 50 (39 - 66) | 70-74 | 53 (39 - 72) |
|  | 75-79 | 48 (38 - 65) | 75-79 | 52 (39 - 69) |
|  | 80-84 | 50 (38 - 65) | 80-84 | 52 (39 - 67) |
|  | 85+ | 51 (39 - 68) | 85+ | 54 (40 - 73) |
| Low-Value Procedure | Overall | 50 (39 - 67) | Overall | 51 (39 - 67) |
|  | 65-69 | 51 (39 - 69) | 65-69 | 52 (39 - 69) |
|  | 70-74 | 50 (38 - 67) | 70-74 | 50 (39 - 67) |
|  | 75-79 | 49 (38 - 65) | 75-79 | 50 (38 - 65) |
|  | 80-84 | 50 (38 - 65) | 80-84 | 50 (38 - 65) |
|  | 85+ | 52 (39 - 69) | 85+ | 51 (39 - 68) |
| PSA Test | Overall | 50 (39 - 67) | Overall | 51 (39 - 68) |
|  | 65-69 | 52 (40 - 69) | 65-69 | 52 (39 - 71) |
|  | 70-74 | 50 (39 - 66) | 70-74 | 51 (39 - 68) |
|  | 75-79 | 48 (38 - 64) | 75-79 | 50 (38 - 66) |
|  | 80-84 | 48 (38 - 62) | 80-84 | 50 (38 - 65) |
|  | 85+ | 48 (38 - 65) | 85+ | 51 (39 - 68) |

Abbreviations: AAA; abdominal aortic aneurysm; ACS: ambulatory care sensitive; DNA: did not attend; GP: general practice; PSA: prostate specific antigen; q1: first quartile; q3: third quartile.

Note: this table does not include unvaccinated individuals. We combined 85-89, 90-94 and 95+ age categories due to low counts.

**Supplementary Table 7. Amongst vaccinated individuals only, median days from index to first COVID-19 vaccination by marker status and age category**

|  | **Presence of marker** | | **Absence of marker** | |
| --- | --- | --- | --- | --- |
| **Marker** | **Age category** | **Median (q1 – q3)** | **Age category** | **Median (q1 – q3)** |
| AAA Screen Males | Overall | 67 (58 - 75) | Overall | 64 (55 - 72) |
|  | 65-69 | 80 (75 - 84) | 65-69 | 80 (74 - 84) |
|  | 70-74 | 71 (66 - 74) | 70-74 | 70 (66 - 74) |
|  | 75-79 | 60 (57 - 66) | 75-79 | 60 (57 - 66) |
|  | 80-84 | 53 (48 - 58) | 80-84 | 53 (47 - 59) |
|  | 85+ | 52 (46 - 58) | 85+ | 53 (47 - 60) |
| Bowel Cancer Screen | Overall | 66 (57 - 74) | Overall | 56 (50 - 65) |
|  | 65-69 | 80 (74 - 84) | 65-69 | 80 (75 - 84) |
|  | 70-74 | 71 (66 - 74) | 70-74 | 71 (66 - 75) |
|  | 75-79 | 60 (57 - 67) | 75-79 | 60 (57 - 67) |
|  | 80-84 | 53 (47 - 59) | 80-84 | 53 (49 - 59) |
|  | 85+ | 52 (46 - 59) | 85+ | 53 (48 - 60) |
| Breast Cancer Screen Females | Overall | 66 (57 - 74) | Overall | 66 (56 - 74) |
|  | 65-69 | 80 (74 - 84) | 65-69 | 80 (74 - 84) |
|  | 70-74 | 71 (66 - 74) | 70-74 | 71 (66 - 74) |
|  | 75-79 | 60 (56 - 66) | 75-79 | 60 (57 - 67) |
|  | 80-84 | 53 (48 - 59) | 80-84 | 53 (48 - 59) |
|  | 85+ | 53 (48 - 59) | 85+ | 53 (48 - 61) |
| Cervical Cancer Screen Females | Overall | 66 (57 - 74) | Overall | 66 (57 - 75) |
|  | 65-69 | 80 (74 - 84) | 65-69 | 80 (74 - 84) |
|  | 70-74 | 71 (66 - 74) | 70-74 | 71 (66 - 74) |
|  | 75-79 | 60 (57 - 66) | 75-79 | 60 (57 - 67) |
|  | 80-84 | 53 (48 - 59) | 80-84 | 53 (48 - 59) |
|  | 85+ | 53 (48 - 60) | 85+ | 53 (48 - 61) |
| NHS Health Checks | Overall | 66 (57 - 74) | Overall | 66 (56 - 74) |
|  | 65-69 | 80 (75 - 84) | 65-69 | 80 (74 - 84) |
|  | 70-74 | 71 (66 - 74) | 70-74 | 71 (66 - 74) |
|  | 75-79 | 60 (57 - 66) | 75-79 | 60 (57 - 67) |
|  | 80-84 | 53 (47 - 59) | 80-84 | 53 (48 - 59) |
|  | 85+ | 53 (49 - 60) | 85+ | 53 (47 - 60) |
| Influenza Vaccination | Overall | 66 (57 - 74) | Overall | 70 (60 - 78) |
|  | 65-69 | 80 (74 - 84) | 65-69 | 80 (76 - 86) |
|  | 70-74 | 70 (66 - 74) | 70-74 | 72 (67 - 76) |
|  | 75-79 | 60 (57 - 66) | 75-79 | 62 (58 - 67) |
|  | 80-84 | 53 (47 - 59) | 80-84 | 55 (50 - 62) |
|  | 85+ | 53 (47 - 60) | 85+ | 56 (50 - 65) |
| Pneumococcal Vaccination | Overall | 66 (57 - 74) | Overall | 70 (60 - 78) |
|  | 65-69 | 79 (74 - 83) | 65-69 | 80 (75 - 85) |
|  | 70-74 | 71 (66 - 74) | 70-74 | 71 (66 - 74) |
|  | 75-79 | 60 (57 - 66) | 75-79 | 61 (57 - 67) |
|  | 80-84 | 53 (47 - 59) | 80-84 | 53 (49 - 60) |
|  | 85+ | 53 (47 - 60) | 85+ | 53 (49 - 62) |
| ACS Hospital Visit | Overall | 66 (57 - 74) | Overall | 67 (58 - 75) |
|  | 65-69 | 79 (72 - 83) | 65-69 | 80 (75 - 84) |
|  | 70-74 | 70 (65 - 74) | 70-74 | 71 (66 - 74) |
|  | 75-79 | 60 (56 - 67) | 75-79 | 60 (57 - 67) |
|  | 80-84 | 53 (49 - 60) | 80-84 | 53 (48 - 59) |
|  | 85+ | 54 (49 - 63) | 85+ | 53 (47 - 60) |
| Blood Pressure Test | Overall | 66 (57 - 74) | Overall | 71 (60 - 78) |
|  | 65-69 | 80 (74 - 84) | 65-69 | 80 (77 - 85) |
|  | 70-74 | 71 (66 - 74) | 70-74 | 72 (67 - 75) |
|  | 75-79 | 60 (57 - 66) | 75-79 | 61 (58 - 67) |
|  | 80-84 | 53 (48 - 59) | 80-84 | 53 (49 - 59) |
|  | 85+ | 53 (47 - 60) | 85+ | 54 (49 - 61) |
| Bone Density Scan | Overall | 66 (57 - 74) | Overall | 66 (57 - 74) |
|  | 65-69 | 79 (73 - 83) | 65-69 | 80 (74 - 84) |
|  | 70-74 | 70 (65 - 74) | 70-74 | 71 (66 - 74) |
|  | 75-79 | 60 (57 - 66) | 75-79 | 60 (57 - 67) |
|  | 80-84 | 53 (48 - 59) | 80-84 | 53 (48 - 59) |
|  | 85+ | 53 (47 - 60) | 85+ | 53 (47 - 60) |
| DNA Primary Care Visit | Overall | 66 (57 - 74) | Overall | 67 (58 - 75) |
|  | 65-69 | 80 (74 - 84) | 65-69 | 80 (75 - 84) |
|  | 70-74 | 71 (66 - 74) | 70-74 | 71 (66 - 74) |
|  | 75-79 | 60 (57 - 67) | 75-79 | 60 (57 - 66) |
|  | 80-84 | 53 (48 - 59) | 80-84 | 53 (47 - 59) |
|  | 85+ | 53 (48 - 61) | 85+ | 53 (47 - 60) |
| GP Visit | Overall | 66 (57 - 74) | Overall | 74 (66 - 81) |
|  | 65-69 | 80 (74 - 84) | 65-69 | 82 (78 - 88) |
|  | 70-74 | 71 (66 - 74) | 70-74 | 73 (68 - 77) |
|  | 75-79 | 60 (57 - 67) | 75-79 | 65 (59 - 70) |
|  | 80-84 | 53 (48 - 59) | 80-84 | 57 (51 - 64) |
|  | 85+ | 53 (47 - 60) | 85+ | 56 (51 - 66) |
| Low-Value Procedure | Overall | 66 (57 - 74) | Overall | 67 (58 - 75) |
|  | 65-69 | 79 (73 - 83) | 65-69 | 80 (75 - 84) |
|  | 70-74 | 70 (65 - 74) | 70-74 | 71 (66 - 74) |
|  | 75-79 | 60 (57 - 66) | 75-79 | 60 (57 - 67) |
|  | 80-84 | 53 (48 - 59) | 80-84 | 53 (48 - 59) |
|  | 85+ | 53 (47 - 61) | 85+ | 53 (47 - 60) |
| PSA Test | Overall | 67 (58 - 75) | Overall | 68 (58 - 76) |
|  | 65-69 | 80 (74 - 83) | 65-69 | 80 (75 - 85) |
|  | 70-74 | 70 (66 - 74) | 70-74 | 71 (66 - 74) |
|  | 75-79 | 60 (57 - 66) | 75-79 | 60 (57 - 67) |
|  | 80-84 | 53 (47 - 58) | 80-84 | 53 (48 - 59) |
|  | 85+ | 53 (46 - 59) | 85+ | 53 (47 - 60) |

Abbreviations: AAA; abdominal aortic aneurysm; ACS: ambulatory care sensitive; DNA: did not attend; GP: general practice; PSA: prostate specific antigen; q1: first quartile; q3: third quartile.

Note: this table does not include unvaccinated individuals and this only includes days until first COVID-19 vaccination. We combined 85-89, 90-94 and 95+ age categories due to low counts.

### Figures

**Supplementary Figure 1. Amongst vaccinated individuals only, box plots for median days from index date to influenza vaccination stratified by marker status and age category**

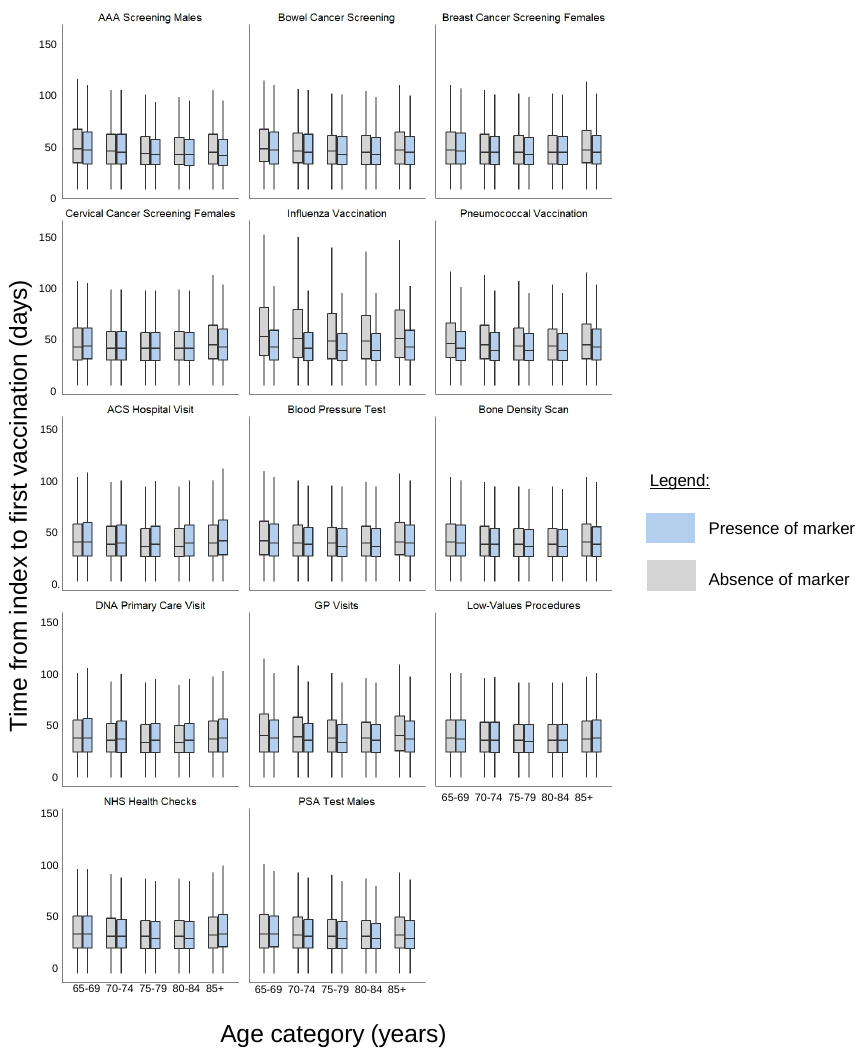

Abbreviations: AAA: abdominal aortic aneurysm; ACS: ambulatory care sensitive; DNA: did not attend; GP: general practice; NHS: national health service; PSA: prostate specific antigen.

Note: the raw data from the figure can be found in **Supplementary Table 6.** This figure does not include unvaccinated individuals. We combined 85-89, 90-94 and 95+ age categories due to low counts.

**Supplementary Figure 2. Amongst vaccinated individuals only, box plots for median days from index to first COVID-19 vaccination stratified by marker status and age category**

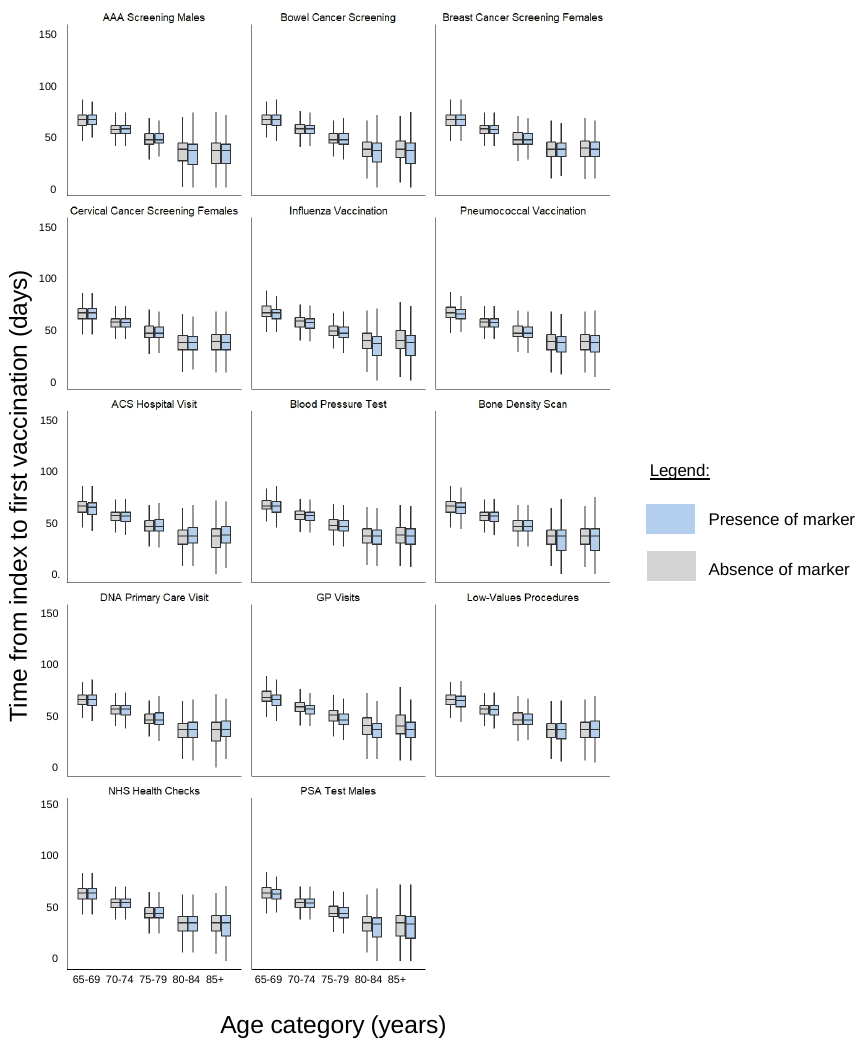

Abbreviations: AAA: abdominal aortic aneurysm; ACS: ambulatory care sensitive; DNA: did not attend; GP: general practice; NHS: national health service; PSA: prostate specific antigen.

Note: the raw data from the figure can be found in **Supplementary Table 7**. We combined 85-89, 90-94 and 95+ age categories due to low counts.
